# Recalculated CNV penetrance: what it does and does not change for counselling

**DOI:** 10.64898/2026.10.06.26364866

**Authors:** Gerome Breen, Harriet Cullen, Heiko Paland, Evangelos Vassos, Laura Kilarski, Mads E. Hauberg, Boris Chaumette, Sergi Papiol, Donna Duffin, Katherine L. Musliner, Wouter J. Peyrot, Eva C. Schulte, Kristiina Tammimies, the ENCAP Group, Kimberley M. Kendall

## Abstract

Goh et al. recently reported that most recurrent copy number variants (CNVs) are far less penetrant for intellectual disability (ID) than previously estimated, and recommended that fourteen should no longer be reported. We show that the lower estimates arise mainly from assuming the population prevalence of ID alone, while case counts derive from broad diagnostic referral cohorts in which about half of individuals were referred for other indications. The proposed formula estimates a risk difference rather than penetrance, and cases and controls were selected, assayed and phenotyped differently. Restricting the outcome to ID also leaves the analysis unable to inform risk for autism, epilepsy, psychiatric, cardiac and other phenotypes. We recommend that families receive phenotype-specific absolute risks for carriers alongside background risks, and that reporting decisions be made locus by locus by guideline bodies, considering all relevant phenotypes. The recalculated estimates inform this debate but are not yet sufficient grounds for changing reporting practice.

## Introduction

We are a collaborative network of clinicians and scientists who contribute to the copy number variant (CNV) evidence base and counsel affected families. Our clinics depend on accurate penetrance figures to inform healthcare. Recently, Goh and colleagues^1^ proposed a modified Bayesian formula to recalculate penetrance for recurrent CNVs. They argue that previous estimates are inflated, and recommend that fourteen CNVs, eleven with recalculated penetrance near 0% for Intellectual Disability (ID), need not be reported.

Re-examining penetrance estimates using population databases is timely, but the proposed revisions raise important methodological concerns. As Goel demonstrated^2^, Goh’s revised metric is an effect measure, thus a rescaled risk difference, and not true carrier penetrance (r_1_). The downward shift in the estimates stems mainly from assuming a lower background disorder prevalence (d = 1.1%), rather than from the formula change. The authors do acknowledge this, reporting that this step produces a three-to twelve-fold reduction^1^. However, the paper’s title, abstract and reporting recommendation are framed around the new definition and formula, and that is how the work is being read and applied. In our experience, some laboratories are already quoting these new estimates in their reports.

We identify four issues: 1) a standard effect measure relabelled as penetrance; 2) the selected prevalence does not correspond to the referral cohorts from which the case data were derived; 3) the exome-based control set is selected, assayed and phenotyped differently from the cases; and 4) the case for non-reporting does not follow from the analyses.

### 1. The new formula is an established effect measure, not conventional penetrance

Conventional penetrance measures the proportion of carriers who exhibit a given phenotype (D). Goh et al.’s formula 2 estimates the proportion of carriers whose phenotype can be attributed to the variant (G), after accounting for background risk (not G).

Let r_1_ = P(D | G) be the absolute risk in carriers and r_0_ = P(D | not G) the risk in non-carriers. Formula 2 is (r_1_ − r_0_)/(1 − r_0_). The attributable fraction among the exposed is AF_e_ = (r_1_ − r_0_)/r_1_, so r_1_ − r_0_ = r_1_ · AF_e_, and substituting:

Formula 2 = (r_1_ · AF_e_)/(1 − r_0_)

Because non-carrier risk is close to the population prevalence, 1 − r_0_ ≈ 1 − d = 0.989 at d = 1.1%, giving

Formula 2 ≈ 1.011 · r_1_ · AF_e_

For the 15q11.2 BP1–BP2 deletion at d = 1.1%, r_1_ = 2.36%, r_0_ = 1.10% and AF_e_ = 0.536, giving 1.28%, published as 1.3%.

Goh et al. state that Formula 2 is distinct from the attributable fraction, and it is: AF_e_ is a proportion of affected carriers, Formula 2 is a proportion of all carriers, and they differ by the factor r_1_/(1 − r_0_), about forty-fold at 15q11.2. But Formula 2 is exactly the risk difference r_1_ − r_0_ = r_1_·AF_e_ rescaled by 1/(1 − r_0_), a factor of 1.011 at d = 1.1%. It is therefore neither AF_e_ nor a new quantity; it is the risk difference, which is a standard effect measure rather than a definition of penetrance^2^.

Consequently, it shares a limitation of excess-risk measures: it captures only the additional cases attributable to the variant, overlooking carriers in whom the variant acts alongside other contributing factors^3^.

### 2. Incompatibility of the Cohort Numerator and Prevalence Baseline

When interpreted as an effect measure, the results are skewed by an assumed prevalence parameter (d = 1.1%) inconsistent with the clinical ascertainment of the cohorts. Goh et al. emphasise that accurately estimating multi-phenotype penetrance requires the clinical referral cohort to mirror the population distribution of those phenotypes. Yet, in one of their primary data sources^4^, ID or developmental delay accounted for only 51% to 54% of recorded indications; the rest include epilepsy, autism without ID, dysmorphic features, and congenital anomalies. When a cohort spans a broad range of neurodevelopmental indications, applying an isolated ID population prevalence (1.1%) rather than the combined prevalence of neurodevelopmental indications (e.g. 5.1%^4^) artificially lowers the calculated risk. The pooled studies themselves used values from 4% to 13%^5^: Rosenfeld et al.^4^ used 5.12% to match their referral series, and Martin et al.^6^ used 13%, the observed proportion of the Geisinger health-system cohort carrying any neurodevelopmental or psychiatric diagnosis (12,039 of 90,595). Figure 1 shows the estimates across that range; at either 5% or 13% the enriched loci sit well above background.

**Figure 1.**
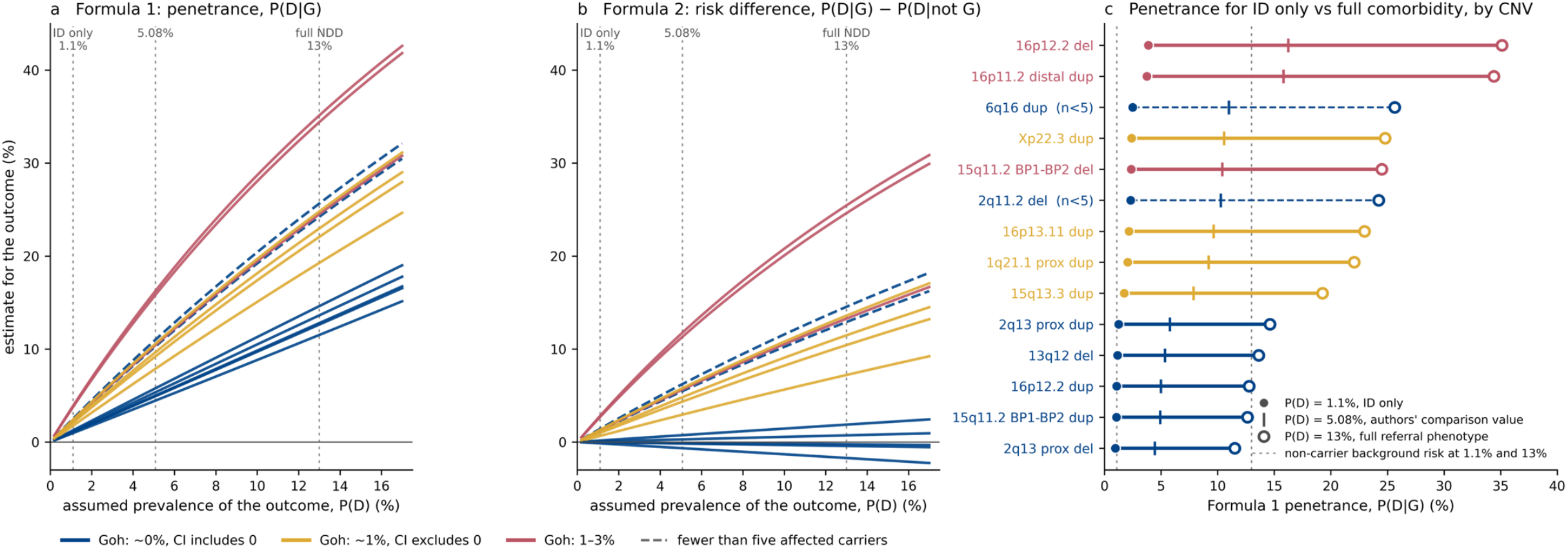
Penetrance depends on the assumed prevalence, which the authors set for ID alone while the case cohorts span the full referral phenotype. Curves and points show the fourteen recurrent CNVs Goh et al. recommend need not be reported, based on the counts in their Table 1; colours are named in (c); dashed lines mark CNVs with fewer than five affected carriers. **(a)** Formula 1, conventional penetrance P(D|G), and **(b)** Formula 2, r_0_ = P(D | not G), against the assumed prevalence of the outcome, with guides at the ID-only prevalence used by the authors (1.1%), their comparison value (5.08%), and a prevalence matching the referral cohorts from which the case counts were drawn (13%). **(c)** Conventional penetrance for each CNV at those three prevalences, with the non-carrier background risk shown as dotted guides. The ranking of CNVs is identical in all three panels and at every prevalence, because both formulas increase monotonically with the case–control carrier ratio; lines in (a) and (b) therefore appear in the same top-to-bottom order as the rows in (c). CNVs with case-control enrichment near 1 sit at the background at any prevalence; the remainder rise from 2–4% to 19–35% when the outcome definition matches the cohort, and their excess over background rises with them. *Note*: Colours are from a palette chosen to remain distinguishable under common forms of colour-vision deficiency and in greyscale (Paul Tol’s high-contrast scheme), and denote the three groups into which Goh et al. place these CNVs; individual CNVs are named in (c).

For example, using the new formula, the penetrance of 1q21.1 distal deletion drops from 38% to 10%, a reduction of 28 percentage points. Changing the assumed prevalence from 5.08% to 1.1%, while keeping the original formula, lowers the estimate to 11%. This shows how little the formula contributes: of the 28-point reduction, 27 points come from applying an ID-only prevalence to a broadly ascertained referral cohort, and only 1 from the formula change.

### 3. Technical Limitations in Exome-Based Control Sets

Goh et al. use gnomAD v4 exomes as controls, the majority of which are from the UK Biobank rather than a diagnostic cohort. The frequencies at the reclassified loci are consistent, within 25%, with array-based calls in the UK Biobank^7^. However, two design issues remain. First, cases are identified with array-based calls spanning the entire interval and undergo clinical curation, whereas controls are identified using exome data from specific exons, with breakpoints inferred and segmental duplications excluded^8^; as a result, the intervals can differ — up to two-fold at 2q13^5^.

Second, the classification of controls. The authors acknowledge that control cohorts may include individuals with ID, biasing estimates downward, and flag the loci in question in Table 1. The larger difficulty is the outcome itself. UK Biobank CNV carriers at 15q11.2, 16p13.11 and 1q21.1^7,9^ exhibit cognitive deficits and higher rates of hypertension, diabetes and mortality ^7,9^, despite having fewer recorded neurodevelopmental diagnoses. Although such individuals are correctly counted as non-cases for an ID-defined outcome, a binary ID threshold registers none of what they experience. The estimate is accurate for the question asked and incomplete as a description of the variant’s effect.

### 4. The case for non-reporting does not follow from the analysis

Four of the fourteen CNVs are enriched in cases strongly enough to pass the authors’ own test of significance: their 95% confidence intervals exclude zero (1q21.1 proximal duplication, 15q13.3 duplication, 16p13.11 duplication and Xp22.3 duplication, each near 1%). By the criterion the paper itself uses, these are positive findings. However, the authors nonetheless discount them, suggesting the small excess in cases may not reflect true penetrance but differences in genetic ancestry between the affected and control cohorts, publication bias, or other confounding. The difficulty is that these factors, if they operate, affect every CNV in the table, not only these four. The same cohorts and the same control set produced the revised estimate for 22q11.2 deletion and moved eleven other CNVs close to zero. A dataset cannot be sound enough to lower those 14 estimates yet unsound enough to explain away a significant signal at four of them.

The recommendation also asserts that these CNVs show little evidence of pathogenicity “for ID or other phenotypes.” The formula estimates risk for a single phenotype, here defined as ID, hence the prevalence estimate of 1.1%. However, carriers referred for epilepsy, autism or congenital anomalies are counted in the numerator, but their outcomes are never measured, so the analysis is silent on every phenotype except the one the denominator was set to. Inferring a lack of pleiotropic effect from an unmeasured outcome conflates absence of evidence with evidence of absence—a logical fallacy the authors themselves acknowledge when noting that CNVs are highly pleiotropic and that their single-phenotype formula cannot capture multi-system risk.

The 16p13.11 duplication illustrates both problems at once. It is one of the four whose recalculated confidence interval excludes zero, so on the authors’ own arithmetic it remains significantly enriched in cases. Independently of cognitive phenotypes, it has also been associated with thoracic aortic aneurysm and dissection^10^. While the need for ongoing aortic surveillance in carriers remains unresolved, reporting such a CNV should depend on further cardiac evidence, not on penetrance figures limited to ID.

## Conclusion

Revisiting recurrent CNV penetrance using population data is long overdue, and we agree with Goh et al. that low-penetrance variants are often over-interpreted. However, the lower estimates largely result from an assumed ID prevalence that does not match the case data, rather than from the new proposed formula. Additionally, differences in how cases and controls are selected and tested further complicate interpretation. The formula itself addresses a different question, giving a standard effect measure rather than conventional penetrance, and via strict prevalence may underestimate a variant’s true impact.

Families may benefit most from knowing the absolute risk for carriers, compared to the background risk in non-carriers, for each phenotype influenced by a CNV, with clear acknowledgement of uncertainty. Guideline bodies should make reporting decisions by considering each locus and all relevant phenotypes. The recalculated risk difference estimates are not penetrance but do offer interesting information for this debate. However, they are not yet sufficient grounds for changing reporting practice.

## Data Availability

This is a response/commentary to a paper in the European Journal of Human Genetics that reanalyses their results.

## European Network for Clinical Application in Psychiatric Genomics (ENCAP) Group

Gerome Breen (King’s College London, London, UK); Boris Chaumette (Université Paris Cité, Paris, France); Harriet Cullen (King’s College London, London, UK); Donna Duffin (Cardiff and Vale University Health Board, Cardiff, UK); Fernando Facal Molina (Servizo Galego de Saúde, Galicia, Spain); Samuelle Fajutrao Falk (Karolinska Institutet, Stockholm, Sweden); Giuseppe Fanelli (Radboud University Medical Center, Nijmegen, The Netherlands); Andreas J. Forstner (University of Bonn, Bonn, Germany); Mads E. Hauberg (Aarhus University, Aarhus, Denmark); Philip R. Jansen (Amsterdam UMC, Amsterdam, The Netherlands); Kimberley M. Kendall (Cardiff University, Cardiff, UK); Laura Kilarski (University of Bonn, Bonn, Germany); Jurjen J. Luykx ([AFFILIATION TO CONFIRM]); Vincent Millischer (Medical University of Vienna, Vienna, Austria); Katherine L. Musliner (Aarhus University Hospital, Aarhus, Denmark); Heiko Paland (Universitätsklinikum Hamburg-Eppendorf, Hamburg, Germany); Sergi Papiol (Max Planck Institute of Psychiatry, Munich, Germany); Wouter J. Peyrot (Amsterdam UMC, Vrije Universiteit, Amsterdam, The Netherlands); Teresa Reynolds de Sousa (Universidade de LisboaUniversity of LisbonInstituto de Saúde Ambiental); Eva C. Schulte (University of Bonn, Bonn, Germany); Kristiina Tammimies (Karolinska Institutet, Stockholm, Sweden); Evangelos Vassos (King’s College London, London, UK); Bjarni Jóhann Vilhjálmsson (National Centre for Register-based Research, Aarhus University, Aarhus, Denmark).

